# How has Covid-19 affected mental health nurses and the delivery of mental health nursing care in the UK? Results of a mixed methods study

**DOI:** 10.1101/2020.11.05.20226472

**Authors:** Una Foye, Christian Dalton-Locke, Jasmine Harju-Seppanen, Rebecca Lane, Lewys Beams, Norha Vera San Juan, Sonia Johnson, Alan Simpson

## Abstract

**Introduction:** While evidence has emerged concerning the impact of Covid-19 on the general population and the challenges facing health services, much less is known regarding how the pandemic has directly affected the delivery of mental health nursing care.

**Aim:** This paper aims to explore how Covid-19 has affected the ability of mental health nurses to deliver care in community and inpatient mental health services in the UK.

**Method:** We investigated staff reports regarding the impact of the Covid-19 pandemic on mental healthcare and mental health service users in the UK, using a mixed methods online survey. A total of 897 nurses across a range of inpatient and community settings participated.

**Discussion:** Key themes within the data explore: new ways of working; remote working; risks of infection/infection control challenges; and the impact on service users. Targeted guidelines are required to support mental health nurses providing care and support during a pandemic to people in severe mental distress, often in unsuitable environments.

**Implications for Practice:** Service developments need to occur alongside tailored guidance and support for staff welfare supported by clear leadership. These findings identify areas requiring attention and investment to prepare for future crises and the consequences of the pandemic.

**Accessible Summary:** *What is known on the subject?:* During the Covid-19 pandemic there has been research considering the impact on medical healthcare professionals and the mental health needs of the general population. However, limited focus has been placed on mental health services or mental health staff providing care in the community and in hospitals. Whilst nurses make up the largest section of the mental health workforce in the UK, the impact that this pandemic has had on their work has been largely ignored.

*What the paper adds to existing knowledge?:* This paper provides a unique insight into the experiences and impact that the Covid-19 pandemic has had on mental health nurses across a range of community and inpatient settings to understand what has changed in their work and the care they can and do provide during this crisis. This includes exploring how services have changed, the move to remote working, the impact of the protective equipment crisis on nurses, and the difficult working conditions facing those in inpatient settings where there is minimal guidance provided.

*What are the implications for practice?:* By understanding the impact the pandemic has had on mental health nursing care, we can understand the gaps in guidance that exist, the challenges being faced, and the impact the crisis has had on care for mental health service users. By doing so we can plan for the ongoing nature of this pandemic as well as the aftermath that the crisis may leave for our service users and workforce alike.

*Relevance Statement:* This paper provides insight into the impact that the Covid-19 pandemic has had on the service and care that mental health nurses are expected to and can provide. As a workforce that often requires ongoing face to face contact with service users, many in serious distress, in inpatient and community settings, it is important that we understand their experiences and the challenges and risks that face this workforce. This will enable us to ensure that future planning, guidance, support and safeguarding can take place during the ongoing and future crises.

## 1. Introduction

The Covid-19 pandemic has placed an increased demand on health and social care services, with major pressures faced by Intensive Care Unit services in general hospitals (*McCabe et al., 2020*). Throughout the pandemic, restrictions placed on individuals, workplaces and services have led to increased concerns about the impact that the pandemic would have in both the short and long-term on mental health services (*Holmes et al., 2020*). A rapid synthesis of published material from the earliest phase of the pandemic (*Sheridan-Rains et al., 2020*) highlighted a broad picture of deteriorating mental health for those with existing difficulties. This review mirrors the emerging picture where much of the empirical literature has placed focus on the impact of the pandemic on individuals (*Fernandez-Aranda et al., 2020; Wang et al., 2020; Qiu et al., 2020; Pierce et al., 2020; Mertens et al., 2020; Mazza et al., 2020; Young Minds, 2020; Mind, 2020; RCPsych, 2020; Cheung, Fong & Bressington, 2020*), with less focus on how it has affected services and the care provided.

Consequently, there is a need for mental health services research to understand the impact of the pandemic and the resulting changing landscape to ensure policymakers, commissioners, and service providers can act to address the needs of those with pre-existing mental health problems, those at risk of developing mental health problems, and the professionals and other staff working in services. This includes the need to focus on the experiences and perspectives of mental health nurses (MHNs) (*O’Connor et al 2020*).

In one of the first studies to focus on the impact of Covid-19 on mental health services from the staff perspective, Johnson and colleagues published results exploring the experiences of 2,180 mental health staff from a range of sectors, professions, and specialties (*Johnson et al., 2020*). Initial results highlight immediate infection control challenges and concerns regarding new ways of working. Multiple rapid adaptations and innovations in response to the crisis were described, especially remote working, which was cautiously welcomed but successful in only some clinical situations. The impact and challenges facing healthcare services are likely to disproportionately affect nursing staff who, in the UK at least, make up the highest number of qualified staff in the mental health workforce (*NHS, 2017*). We are already seeing the negative impact that Covid-19 has had on nurses in ICUs, COVID-19 designated hospitals, and departments involved with treating COVID-19 patients, with these nurses reporting higher scores in mental health outcomes (Chen et al., 2020).

The crisis has exacerbated the longstanding problems faced by nurses in relation to inequalities, inadequate working conditions and chronic excessive work pressures, exemplified by the increasing vacancies, absenteeism, turnover and intentions to quit reported prior to the pandemic, with staff having reported feeling ‘broken’, ‘exhausted’ and ‘on their knees’ (The Kings Fund, 2020). Beyond issues related to workload and stress, the sobering impact of the pandemic can be seen with the cost to life that has been seen within this workforce, with many nursing staff, including mental health nurses, working on the frontline during Covid-19 having lost their lives (RCNi, 2020).

Mental health nurses are the staff group most likely to be involved in face-to-face interactions with patients in inpatient settings who may be highly distressed and/or frustrated at the restrictions typically imposed that include the use of rules and procedures to maintain safety, enforced medication, seclusion and restraint (*Bowers et al 2013*), even before further pandemic-related impositions. Additionally, a high proportion of admissions to mental health wards are now enforced detentions under the Mental Health Act, further fuelling the potential for conflict and use of containment measures (*Akther et al, 2019*).

Consequently, this paper provides a unique and more detailed in-depth analysis of the responses from mental health nurses that responded to the *Johnson et al (2020)* survey. To our knowledge this is the first work that has highlighted the experiences and needs of MHNs during the pandemic.

### 1.2. Aim

The aim of this study is to explore how Covid-19 has affected the mental health nurse workforce in order to understand what changes to services have been made and how these affect the care and treatment that MHNs provide across a range of mental health settings.

## 2. Methodology

The findings reported in this study were drawn from data collected from a survey exploring the perspectives and experiences of staff working in mental health inpatient, community and specialist services across the UK during the early stages of the national Covid-19 related restrictions, from 22 April 2020 to 12 May 2020. The university research ethics committee approved this study *<u>[ethics information to be added here</u>]*. The survey included both structured and open-ended questions and followed a branching system allowing for sections of questions relating to specific settings (e.g. inpatient or community) and specialities (e.g., sections dedicated for staff working in perinatal settings). Given the lack of literature regarding the specific MHN experiences, we conducted a secondary, more detailed analysis of this survey data using the responses provided by participants describing themselves as nurses.

### 2.1. Questionnaire design

The questionnaire was developed to include questions covering a range of sections to address staff experiences during Covid-19. These sections followed three main sets of questions asked of all participants, covering: (1) Challenges at work during the Covid-19 pandemic, (2) Problems currently faced by mental health service users and family carers (from a staff perspective), and (3) Sources of help at work in managing the impact of the pandemic. The survey took around 15 to 30 minutes for participants to complete, depending on the number of open-ended questions answered and the depth of answer provided (*Johnson et al., 2020*). The study used an explanatory mixed methods design, a methodological design consisting of two distinct phases: quantitative followed by qualitative (*Creswell et al. 2003*). The qualitative data are collected and analysed second in the sequence and help elaborate on the quantitative results.

Responses to quantitative questions were scored on a four-point Likert scale of relevancy ranging from ‘slightly’ to ‘extremely’ relevant, with additional options of not relevant or not applicable. The inclusion of a range of open-ended qualitative questions, that were incorporated throughout the questionnaire, allowed participants to add details to the quantitative responses. Further details regarding the development of the survey and recruitment can be found in the publication describing the whole dataset (*Johnson et al., 2020*).

Participants were presented with all questions, both qualitative and quantitative, with no questions being mandatory. Demographic questions were at the end of the questionnaire meaning that fewer participants completed this section than earlier questions related to their practice.

### 2.2. Analysis

All data for participants who identified as a MHN were extracted to Microsoft Excel. Quantitative data were then imported to SPSS 26 for analysis (IMB, 2019). The quantitative data were used to produce descriptive statistics to summarise relevant aspects of the responses. Participants were not required to answer all questions therefore descriptive percentages were calculated based on the total number of participants who provided an answer to that question rather than from the full sample.

The qualitative data was analysed within Excel, using a thematic analytic approach, as outlined by Braun and Clarke (*2006*). Team members read and re-read the data noting aspects of interest. Following this, a list of codes was generated. Microsoft Excel was used to record the extracted chunks of data and associated codes. Following initial coding, related codes were sorted, grouped and labelled as preliminary themes, defined as the central concepts that capture and summarize the core point of a coherent and meaningful pattern in the data (*Braun & Clarke, 2006*).

Participants responded to a range of the quantitative and qualitative items. As questions were not mandatory the number of participants responding to each item varied across the survey meaning that denominations presented within the results represent those who answered each item.

## 3. Results

### 3.1. Description of sample

A total of 897 MHNs were included within the dataset. Most of the sample worked in the National Health Service (NHS) (n=870, 97%). Only a small proportion of participants worked outside of the NHS with 13 working in the private sector, six in community user-led organisations, five in the voluntary sector and three in local government. The majority worked with adults (n=622, 69.8%) and older adults (n=369, 41.4%), while a smaller proportion of the sample worked with children and younger people (n=149, 16.7%). The settings and specialisms are outlined in Table 1 and Table 2 outlines the demographics of the sample.

**Table 1:** Participants by Sector and Specialism*, **

| **Sector/ Specialism** | **Setting** | **N** | **Percentage of sample** |
| --- | --- | --- | --- |
| Inpatient | Inpatient Wards | 333 | 37.40% |
| Inpatient | Crisis Housing | 18 | 2.00% |
| Inpatient | Residential Services | 12 | 1.30% |
| Community | Crisis Services | 176 | 19.80% |
| Community | Community Mental Health Team (CMHT) | 404 | 45.30% |
| Community | Community Groups | 32 | 3.60% |
| Community | Other | 96 | 10.80% |
| Adult Services |  | 622 | 69.8% |
| Older Adult Services |  | 369 | 41.4% |
| Work with children and young people |  | 149 | 16.7% |
| Intellectual Disabilities |  | 241 | 27.00% |
| Forensic Services |  | 180 | 20.20% |
| Perinatal |  | 158 | 17.70% |
| Drugs & Alcohol Services |  | 219 | 24.60% |
| Eating Disorders |  | 186 | 20.90% |

*\* Participants may work across more than one sector (e.g., NHS and voluntary), in more than one setting (e.g., an inpatient service and crisis assessment service) and with more than one patient group (e.g., working age adults and forensic). Percentages for these variables therefore do not add to 100%.*
*\*\* Percentages are of participants that provided an answer*

**Table 2.** Demographics of the sample, with breakdown for setting

|  |  | Total Sample | Inpatient Sample | Community Sample |
| --- | --- | --- | --- | --- |
| **Gender** | **Male** | 119 (22%) | 43 (31.8%) | 88 (20.7) |
|  | **Female** | 421 (77.9%) | 154 (77.7%) | 337 (79.3%) |
|  | **Other** | 1 (0.1%) | 1 (0.5%) | 0 (0%) |
| **Age** | **Under 25** | 17 (3.1%) | 14 (7%) | 4 (0.9%) |
|  | **25 – 34** | 103 (19%) | 45 (22.6%) | 68 (15.9%) |
|  | **35 – 44** | 131 (24.1%) | 43 (21.6%) | 108 (25.3%) |
|  | **45 – 54** | 191 (35.2%) | 61 (30.7%) | 158 (37%) |
|  | **55 – 64** | 99 (18.2%) | 34 (17.1%) | 88 (20.6%) |
|  | **65+** | 2 (0.4%) | 2 (1%) | 1 (0.2%) |
| **Ethnicity** | **White** | 464 (88.5%) | 165 (86.4%) | 373 (90.3%) |
|  | **Asian** | 9 (1.7%) | 3 (1.6%) | 6 (1.5%) |
|  | **Black** | 30 (5.7%) | 17 (8.9%) | 16 (3.9%) |
|  | **Mixed/ Multiple Ethnic groups** | 15 (2.9%) | 5 (2.6%) | 13 (3.1%) |
|  | **Other/ Prefer not to say** | 6 (1.1%) | 1 (0.5%) | 5 (1.2%) |
| **Caring Role** | **Children under 18** | 203 (37%) | 32.5% (n=65) | 39.39% (n=169) |
|  | **Other** | 164 (30.2%) | 54 (27.1%) | 139 (32.5%) |
| **Working Situation** | **Mainly workplace** | 294 (53.6%) | 166 (82.6%) | 166 (38.5%) |
|  | **Mix of workplace and working from home** | 164 (29.9%) | 25 (12.4%) | 171 (39.7%) |
|  | **Working from home** | 13.5% (n=74) | 2% (n=4) | 19.3% (n=83) |
|  | **Sick or self-isolating/ shielding** | 3% (n=16) | 3% (n=6) | 2.5% (n=11) |
| **Had Covid-19** | **Yes, confirmed** | 3.3% (n=18) | 7% (n=14) | 1.86% (n=8) |
|  | **Yes, suspected** | 23.8% (n=130) | 23.5% (n=47) | 24.9% (n=107) |
|  | **No** | 72.9% (n=398) | 69.5% (n=139) | 73.19% (n=314) |

\* Participants may work across more than one sector (e.g., NHS and voluntary), in more than one setting (e.g., an inpatient service and crisis assessment service) and with more than one patient group (e.g., working age adults and forensic). Percentages for these variables therefore do not add to 100%.
\*\* Percentages are of participants that provided an answer

The top concerns that participants identified as very or extremely relevant to their work are outlined in Table 3 below, with Tables 4 and 5 outlining the differences in concerns of those working in community and inpatient settings.

**Table 3.** Top Concerns Mental Health Nurses report in relation to the impact of Covid-19 *, **

| **Top Concerns (all mental health nurses)** |  |  |
| --- | --- | --- |
| **Concern type** | **N** | **Percentage** |
| 'Having to adapt too quickly to new ways of working' | 478 | 61.4% |
| 'The risk I could be infected at work' | 412 | 53.5% |
| 'Pressures for the need to support colleagues' | 362 | 47.0% |
| 'The risk family and friends may be infected through me' | 365 | 47.0% |
| 'The risk Covid-19 will spread between service users' | 342 | 44.0% |
| 'Having to respond to additional mental health needs that appear to result from Covid-19' | 333 | 42.9% |
| 'Service users no longer getting an acceptable service due to service reconfiguration due to Covid-19' | 311 | 40.0% |
| 'Having to learn to use new technology too quickly' | 304 | 39.6% |
| 'Problems from lack of testing' | 312 | 38.4% |
| 'Concern that physical health care received by service users I work with may not be adequate' | 296 | 38.0% |

\* Numbers are calculated based on respondents answering Extremely or very relevant with missing removed meaning that concerns with higher N responses may have a lower percentage.
\*\* Percentages are of participants that provided an answer

**Table 4.** Top Concerns Inpatient Mental Health Nurses report in relation to the impact of Covid-19 *, **

| **Top Concerns for inpatient mental health nurses** |  |  |
| --- | --- | --- |
| **Concern type** | **N** | **Percentage** |
| 'The risk I could be infected at work' | 213 | 67.8% |
| 'The risk Covid-19 will spread between service users' | 208 | 66.2% |
| 'Having to adapt too quickly to new ways of working' | 205 | 64.3% |
| 'The risk family and friends may be infected through me' | 189 | 60.2% |
| 'Pressures for the need to support colleagues' | 177 | 56.7% |
| 'Difficult putting infection control into practice' | 156 | 50.0% |
| 'Greater workload than usual' | 140 | 44.6% |
| 'Having to respond to additional mental health needs' | 137 | 43.6% |
| 'Problems from lack of testing' | 136 | 43.5% |
| 'Concern that physical health care received by service users' | 120 | 38.3% |

*\* Participants may work across more than one sector (e.g., NHS and voluntary), in more than one setting (e.g., an inpatient service and crisis assessment service) and with more than one patient group (e.g., working age adults and forensic). Percentages for these variables therefore do not add to 100%.*
**\*\* Percentages are of participants that provided an answer**

**Table 5.** Top Concerns Community Mental Health Nurses report in relation to the impact of Covid-19 *, **

| **Top Concerns for community mental health nurses** |  |  |
| --- | --- | --- |
| **Concern type** | **N** | **Percentage** |
| 'Having to adapt too quickly to new ways of working' | 363 | 58.0% |
| 'The risk I could be infected at work' | 293 | 47.1% |
| 'Pressures for the need to support colleagues' | 272 | 44.0% |
| 'Having to respond to additional mental health needs' | 259 | 41.6% |
| 'The risk family and friends may be infected through me' | 257 | 41.2% |
| 'Having to learn to use new technology too quickly' | 254 | 40.8% |
| 'Service users no longer getting an acceptable service due to service reconfiguration' | 243 | 40.4% |
| 'Being expected to use new technologies without reliable access' | 230 | 36.9% |
| 'Concern that physical health care received by service users' | 224 | 36.0% |
| 'The risk Covid-19 will spread between service users' | 194 | 32.7% |

*\* Participants may work across more than one sector (e.g., NHS and voluntary), in more than one setting (e.g., an inpatient service and crisis assessment service) and with more than one patient group (e.g., working age adults and forensic). Percentages for these variables therefore do not add to 100%.*
*\*\* Percentages are of participants that provided an answer*

### 3.2. Impact on working

#### 3.2.1. New ways of working

As outlined in Table 3, the top concern for MHN participants was the quick adaption that many had to make to their way of working, with over 60% of all participants feeling this was too quick. This was the top concern for community staff who largely had to adapt to working remotely and providing services via the telephone and new video and online platforms. Many reported that using such new technologies was hampered by logistical challenges such as poor internet connection, hardware availability and lack of privacy to conduct appointments.

> *“Managing remote working, video conferencing and online meetings with dated equipment.” (Perinatal service, London)*

This move to remote appointments and care had mixed results with some participants noting it as a useful method of ensuring continuity of care. In contrast, others felt that this method did not lend itself well to some populations for a range of reasons including access, logistics and lack of trust:

> *“Remote appointments work well with some clients but they have to a) be well enough to use the tech e.g., not feel their device is being monitored b) they have to have the tech in the first place, lots of our clients do not have smart phones. Some clients report that it is an invasion of privacy.” (Adult Crisis service, London)*

Difficulties engaging service users with remote appointments was the most frequently reported challenge, particularly with those who are experiencing cognitive impairments, young people, those with autism or experiencing psychosis and/or paranoia due to levels of understanding, access and trust in the technology used. The result of these difficulties engaging service users means that assessments or appointments are hampered:

> *“Remote video appointments are almost impossible with my clients who are being assessed for or have dementia as many do not have or understand the technology.” (Older Adults, Scotland)*
>
> *“Very challenging with a psychosis client group who are often suspicious of technology / anxious about using it.” (1-2-1 service, South West England)*

In addition, remote means of working also impacts on ways that teams can work. While many welcomed the move to the use of online platforms such as Attend Anywhere, Zoom and Microsoft Teams for meetings, challenges were reported when trying to implement care plans in this way, particularly in relation to multidisciplinary team meetings;

> *“Remote MDT [multi-disciplinary team] working has led to poor implementation of changes in treatment plans.” (Older Adults, South West England)*

Further to these issues, participants noted that this way of working was exhausting as it often meant that they were more pressured to have back-to-back appointments and thus have more meetings or appointments in one day than previously expected. This added to workload pressures and feelings of burnout among staff:

> *“Staff have reported remote appointments as very intense & although saving on travel recognising they usually use the travel time as down time/mental process time before the next patient.” (Perinatal service, East Midlands)*

#### 3.2.2. Increased workload

When asked how Covid-19 impacted on people’s working, just over a quarter of participants who answered this question felt that they were working longer hours as a direct result of the pandemic (n=203 of 779, 26.1%). This was higher in those working in inpatient settings (n=118 of 315, 37.5%) compared to those working in the community (n=157 of 624, 25.2%). Overall, around a third of participants felt that their workload had increased as a result of Covid-19 (n=270 of 779, 34.7%) and again this was higher in the inpatient groups (n=138 of 314, 44%) compared to community nursing (n=204 of 626, 32.6%). Overall, pressures resulting from staff shortages were reported by over a quarter (n=209 of 769, 27.2%).

### 3.3. Impact on service users

The impact of changes was reported to be a considerable worry for MHNs in relation to how this influenced the care provided to service users, with 40% of the sample (n=331 of 776) feeling that the statement ‘service users no longer getting an acceptable service due to service reconfiguration’ was very or extremely relevant to their work. This was one of the top concerns that MHNs had regarding the impact of Covid-19 on themselves and their work (*see Table 3*). Over half of the sample (n=394 of 736, 53.5%) were concerned about the lack of access to usual support from NHS mental health services for service users.

As service users with severe mental illness (SMI) have higher comorbid physical health conditions, several participants raised concerns over the reduction in services to support these additional medical needs or fear of attending services for physical health needs:

> *“[increased concern for those] not accessing physical health care, not wishing to burden services including A and E.” (Community Service Manager, North East England)*

Twenty percent (n=150 of 731) of the sample noted concern regarding service users’ access to and administration of medication:

> *“Numerous admissions from people who are not being fully supported in community, lack of CPN [community psychiatric nurse] visits. Depot injections being extended. This is leading to admissions.” (Adult Inpatient Service, Scotland)*

### 3.4. Risk of infection

A prominent theme within the data was the concern that the MHN workforce felt in relation to the risk of Covid-19 infection. This was the second leading concern for participants across all sectors (e.g., inpatient and community settings) (see Table 3), with over half of participants reporting they were worried they could be infected in work. This concern regarding risk was the top concern for inpatient MHNs with two-thirds (67.8%) of inpatient nurses reporting this was very or extremely relevant to them (see Table 4), with risk to service users and family or friends also being a concern for over 60% of the inpatient MHNs. Such level of concern was lower in the community MHN sample with 47.1% reporting concern for their own risk and 41% regarding concern for risk to family or friends. Considerably lower levels of concern were reported in relation to risk to service users (see Table 5).

#### 3.4.1. The work environment

Across both the community and inpatient settings, MHNs reported that the work environment made it incredibly difficult to follow social distancing guidelines or infection control measures effectively. When asked ‘Is it practical to follow consistently the rules you have been given on infection control at work?’, almost half of participants (n=270 of 609, 44.3%) answered no. Both the ward layout and office spaces posed considerable issues. Those working in the community and using office space noted that the layout of buildings, lack of computers and space meant it was *‘impossible to keep 2 meters distance’* from colleagues as the space was too small and too congested.

> *“The environment doesn’t allow the guidance to be followed.” (Adult Inpatient Service, North East)*

Within inpatient settings these challenges were more pronounced with shared rooms and ward layout meaning social distancing was unachievable or difficult and that the environment was *‘impractical for proper infection control’*.

> *“Self-isolation - none of the rooms are ensuite so young people that are [Covid-19] positive and isolating will need to exit their room, enter the corridor briefly to then use their designated toilet. Social distancing - nursing office is too small to facilitate this and there aren’t enough working computers to have one nursing staff in each office. Any restraint/debrief means we break social distancing.” (CAMHS Inpatient Service, London)*

#### 3.4.2. The challenges with Personal Protective Equipment (PPE)

The lack of PPE was cited by many as a significant issue. A quarter of participants (198 of 778, 25.4%) reported that they felt there was a lack of PPE needed for infection control. While some participants noted that their management had gone above and beyond to provide sufficient PPE, many noted there was a lack of suitable PPE available to them for their work. One participant directly linked this to them being infected:

> *“PPE provided has been very inadequate and, I believe, the cause of me getting infected by a patient.” (Forensic Inpatient Service, South East England)*

Many participants felt that the downgrading and rewriting of PPE guidelines, and sometimes conflicting advice caused confusion and difficulties for staff on the frontline:

> *“incongruent advice on which PPE to use and the downgrading that left nurses confused and not wearing gloves at all except in personal care!” (Older Adults Inpatient Services, South East England)*

While challenges were reported regarding access and use of PPE in these settings, many MHNs also noted that they found it helpful when they were provided with refresher training in infection control and shown how to use it appropriately.

Within the community there were additional challenges surrounding the topic of PPE particularly in relation to home visits and challenges with donning and doffing PPE. For some this was due to the inadequate guidelines regarding safe methods to put on, take off and dispose of PPE when doing home visits:

> *“I was not told how to dispose of PPE in the community and have been given different advice (none of it written). I have not been able to ‘don and doff’ in people’s homes.” (CMHT, South East England)*

Practical issues were considered a considerable challenge for community MHNs who reported that the nature of their work meant having a lack of basic facilities for working with PPE that are often assumed, meaning guidance and allowances for other means of working had not been considered;

> *“PPE as clinical waste and prolonged driving hours in order to be able to dispose of this safely.” (Perinatal Service, North East)*

Within inpatient settings there was a marked challenge for infection control raised in relation to emergency situations, such as restraints, where there may not be time to don PPE or where wearing PPE may present a risk;

> *“Can’t always put PPE when responding to patient ligaturing.” (Adult Inpatient Services, North West England)*

In one case a participant highlighted that the nature of mental health nursing work meant that additional risks existed that PPE guidance would not pick up but was salient for guidance to consider when understanding the wider risks and necessary precautions:

> *“The children often display risk behaviours, suicidal ideation and self-harm and have tried to take PPE from staff in order to self-suffocate and choke themselves.” (CAMHS Inpatient Service, North East England)*

#### 3.4.3. Service users’ wellness and understanding

In addition to the challenges facing MHNs directly, many noted that the nature of working in mental health services, particularly where there are high levels of acuity, meant that social distancing and adhering to guidelines was difficult even where PPE was available and space was provided, due to service users being mentally unwell. Almost half of the sample (n=359 of 727, 49.4%) reported concern that service users may have difficulties understanding or following current government requirements on social distancing, self-isolation and/or shielding. Staff reported that in many cases service users may lack trust in the services or have diminished cognition and this may explain the challenges that MHNs on wards experienced in relation to problems with social distancing and adhering to guidelines:

> *“Clients with psychosis who have delusional frameworks around government conspiracy/being infected.” (CMHT, London)*
>
> *“Those whose mental state prevents them from understanding the pandemic and infection control risk therefore making them more likely to spread the virus/become infected.” (Inpatient Forensic Service, London)*

### 3.5. Sources of support

When asked about the main sources of support that MHNs felt were very or extremely useful to them during the pandemic, 70% of MHNs said the support and advice from managers was key. In particular, the guidance regarding clinical issues developed by managers and local leadership within mental health settings was important to staff navigating the changing landscape. This helps MHNs to overcome some of the key challenges identified, namely the frequently changing and confusing nature of guidance and its lack of specificity to mental health settings that bring perhaps unique challenges. More generic sources of support via the sources such as the media or voluntary groups were felt to be less useful for staff than these more MHN-led sources.

## 4. Discussion

To the best of our knowledge, this is the first paper to report in detail the experiences of mental health nursing staff during the Covid-19 pandemic. While many of the issues identified reflect similar themes reported within the original study of the wider mental health workforce (*Johnson et al., 2020*), the analysis presented in this paper shows the key and often unique challenges and concerns that face MHNs. The findings highlight the major challenges facing inpatient MHNs in providing care and treatment while wearing PPE and maintaining social distancing with patients who may not have capacity to understand guidelines, and within environments that are often unsuitable and unadaptable. Furthermore, this paper highlights the added complexity of providing care safely on wards where there are potential suicide risks and the need to implement restrictive practices when required. This unique complexity is generally missing from the wider literature and unlikely to be addressed within more generic guidelines. Whilst many of the challenges facing community based MHNs are reflected in the wider professionals’ experiences of rapidly moving to remote working, many MHNs continued to provide care, treatment and support in people’s homes in the community, often in challenging environments. Throughout, the guidance for these MHNs was reportedly lacking and needs addressing swiftly to reduce the risks that are placed on the workforce and those they are caring for.

The response to the Covid-19 crisis has irrefutability had a significant impact on mental health services (*Moreno et al., 2020; Marshall, Bibby & Abbs, 2020*). As we have seen in the emerging literature surrounding this topic, MHNs are experiencing similar work stresses as other healthcare professions in relation to changes to workload and moves towards the use of remote technologies (*Sheridan-Rains et al., 2020; Nuffield Trust, 2020*). Many of these shifts in clinical practice, especially the greater use of technologies and remote working, are likely to remain. Therefore, the commissioning of services, pre-registration nursing education and the continuing professional development of MHNs will need to consider and evolve in line with the emerging lessons of the pandemic. Additionally, the need for reliable hardware, internet connection and support including appropriate training is necessary in ensuring that care can be provided on these platforms in the uninterrupted manner that face-to-face sessions require. As shown in the findings, MHNs are concerned over increased remote appointments in circumstances where this means that some people may not get seen, such as those without access to internet or hardware and people with psychosis experiencing paranoia. It is therefore important to research and understand the impact of remote appointments on therapeutic relationships with service users, particularly with those where it is not easy or perhaps appropriate.

These issues also need to be considered to ensure that the socioeconomic inequalities already facing service users regarding access to services are not widened by requiring laptops, stable wi-fi connections, mobile data and housing with privacy, which are currently considered a luxury rather than a right. The findings presented in this paper also draws attention to the wider inequalities that face mental health service users. Patients from Black, Asian or other minority ethnic communities are likely to be disproportionately represented in our inpatient wards (*Barnett et al., 2019*) and those with severe mental illness are more likely to develop physical health comorbidities such as diabetes, cardiovascular, and respiratory illnesses (*Reeves et al., 2018*). All of these are linked to a higher risk of having severe outcomes in those who contract Covid-19 (*Aldridge et al., 2020; Clark et al., 2020, Wang et al 2020*). Hospitals are noted as hotspots for people being infected with Covid-19. These risks may be due to some of the challenges outlined within this paper, such as difficulties adhering to social distancing or donning PPE in cramped environments, meaning these populations are at a considerable additional risk. Consequently, the difficulties noted within this paper as experienced by staff within these settings must be considered and acted on to ensure the safety of this vulnerable patient population and the staff that care for them.

Shortages of PPE have been widely reported within other healthcare environments (*Kamerow, 2020; Livingston et al., 2020*), but this paper identifies the specialist issues concerning the use of PPE within mental healthcare that have not been at the forefront of the debate. Issues related to donning and doffing PPE when undertaking home visits, disposal issues and the challenges of wearing PPE on wards with patients at risk of suicide and self-harm have all been highlighted within this paper and require further attention, guidance and support for nursing staff trying to manage these challenges and to keep patients and staff safe. Confusion and a lack of targeted guidance for MHNs working in the community regarding PPE donning and doffing, and for those in inpatient settings regarding the management of social distancing with mentally unwell patients and high-risk restraint challenges are among the specialist areas that require attention to ensure MHNs and service users remain safe during further spikes and viral outbreaks. As there is a need to be responsive in such situations due to risk, there is not always time for MHNs, who are almost uniquely the staff called upon to act in these situations, to implement infection control strategies such as washing their hands or putting on all the PPE during psychiatric or medical emergencies. As most other professions are not directly involved in restraint, such guidance for the management of these situations highlights the need for nursing leadership to ensure that nursing specific issues like this are picked up and addressed to tackle hospital-based hotspots. It may also require some creative thinking and perhaps greater engagement with approaches aimed at preventing or deescalating conflicts on wards (*Cole, 2020*).

Even where PPE has been available and both staff and service users are following guidance to socially distance, the environments in which staff are working appears to impinge on their ability to do so. This was salient for wards which have been noted for years as being out of date and posed challenges for safety in relation to ligature protocols (*CQC, 2017*). This new crisis adds to the evidence that many buildings and wards in which care is being provided are no longer fit for purpose when facing new crises and challenges and highlights the need for investment and environmental planning within our mental health services.

## 5. Limitations and future directions

While the study allows initial insights into how Covid-19 has impacted on mental health nursing, there are limitations to consider. Firstly, it must be noted that this data presents only staff perspectives. There is a need to consider questions regarding the impact on care from the service user and carer perspectives to fully understand the impact Covid-19 has had on mental health services. In addition, a key limitation is that the study numbers across groups are not similar or matched meaning that the study lacked power to undertake comparative statistical analysis. There is a need to address the homogeneous nature of the sample. While recruitment efforts were made to target Black, Asian and ethnic minority (BAME) populations, the sample remains lacking in diversity thus findings are skewed to represent a white sample rather than acknowledging specific risks and issues that may be in play for BAME colleagues.

Furthermore, it must be highlighted that the data was taken from a wider national study of all healthcare professionals and there were no nursing specific questions within the survey. The findings highlighting the need for specialist nursing information and guidance points towards the need to conduct research specifically designed for our mental health nursing colleagues’ experiences. We hope that this paper will start the conversation towards designing nurse-led research within this area to address aspects that more general mental health research may be missing.

Future research should consider a longitudinal perspective on this pandemic, using repeated data points to see if and how experiences have or have not changed over the course of the pandemic and in the long term.

## 6. Conclusion

The findings have shown key areas that require attention and guidance during the ongoing Covid-19 crisis. This includes the challenges of the swift shift to remote working in community services, ensuring the safe use of protective clothing in people’s homes or on cramped wards, and the need for targeted guidance and support for MHNs working in inpatient settings, where they are faced with unique challenges in maintaining social distancing and interacting with mentally distressed and confused patients. These findings show that mental health nurses face unique challenges and are impacted differently across different sectors. Covid-19 is not necessarily a shared experience; therefore, research needs to provide a more focused lens on individual settings, e.g., inpatient wards, child and adolescent services, older people’s service, eating disorder units, and crisis and assertive outreach services, to ensure these are managed effectively. This paper touches on some of the current demands and challenges identified, but we also need to consider how much and how fast the system will need to change if we see a rapid escalation of mental health presentations over the next year as a consequence of the pandemic, and how mental health nursing staff will be supported or prepared for this.

## Data Availability

The datasets generated during and/or analysed during the current study are available from the corresponding author on reasonable request

